# Initial proteinuria reduction and adverse kidney outcomes in IgA nephropathy: An analysis from the J-IGACS

**DOI:** 10.1101/2025.06.15.25329622

**Authors:** Takaya Sasaki, Nobuo Tsuboi, Kentaro Koike, Hiroyuki Ueda, Masahiro Okabe, Shinya Yokote, Akihiro Shimizu, Keita Hirano, Tetsuya Kawamura, Takashi Yokoo, Yusuke Suzuki, the J-IGACS working group

**Affiliations:** Division of Nephrology and Hypertension, Department of Internal Medicine, The Jikei University School of Medicine, Tokyo, Japan; Department of Nephrology, Juntendo University Faculty of Medicine, Tokyo, Japan

**Author notes:** Correspondence to: Nobuo Tsuboi, M.D., Ph.D. Division of Nephrology and Hypertension, Department of Internal Medicine, The Jikei University School of Medicine, 3-25-8 Nishi-Shimbashi, Minato-ku, Tokyo, Japan.

**Keywords:** IgA nephropathy, proteinuria, eGFR slope, kidney outcome

## Abstract

**Background:** Proteinuria reduction is considered a potential surrogate endpoint predictive of reflecting long-term kidney prognosis in IgA nephropathy (IgAN). However, its association with adverse kidney outcomes and IgAN-related decline in estimated glomerular filtration rate (eGFR) remains uncertain.

**Methods:** Patients with biopsy-proven IgAN from the Japan IgA Nephropathy Cohort Study (J-IGACS) were analyzed. Participants were categorized into tertiles based on their 12-month proteinuria-to-baseline proteinuria ratios. The primary outcome was a composite of ≥40% eGFR decline or initiation of kidney-replacement therapy. Associations between proteinuria ratio and outcomes were assessed using Cox proportional hazards models and restricted cubic splines. Multivariable analyses adjusted for age, sex, baseline eGFR, log-transformed proteinuria, Oxford classification scores, and use of corticosteroids and renin-angiotensin-aldosterone system inhibitors within 12 months.

**Results:** Among 793 patients, those in the lowest tertile (greatest proteinuria reduction) had significantly lower risk of the primary endpoint (P for trend <0.001) and a more favorable eGFR slope. Spline analysis showed a continuous, dose-response association between proteinuria ratio and improved outcomes. These findings remained robust in sensitivity analyses restricted to patients likely qualifying for clinical trials. The results showed that patients with lower proteinuria ratios tended to have slower rates of eGFR decline (P for trend <0.001), even after multivariable adjustment.

**Conclusion:** Proteinuria reduction within the first-year post-diagnosis is independently associated with lower risk of adverse kidney outcomes and a slower decline in kidney function in patients with IgAN. These results support the use of proteinuria reduction as a surrogate endpoint in both clinical trials and disease management.

## Introduction

Immunoglobulin A nephropathy (IgAN) is the most prevalent primary glomerulonephritis globally and a major contributor to progressive chronic kidney disease (CKD) and kidney failure [1–4]. Despite advances in understanding its pathophysiology and the development of histopathologic classification systems such as the Oxford Classification [5, 6], treatment strategies remain largely supportive, with limited disease-specific therapies approved to date.

Proteinuria is a well-established risk factor in IgAN and is frequently utilized in clinical practice as a target for therapeutic intervention [7, 8]. Several studies have proposed proteinuria reduction as a potential surrogate endpoint for kidney outcomes, particularly in the context of clinical trials [9, 10]. However, while short-term reduction in proteinuria is generally considered favorable, the extent to which this improvement translates into long-term preservation of kidney function, particularly as reflected by the slope of the estimated glomerular filtration rate (eGFR), remains incompletely elucidated, especially in Asian countries, including Japan, where high-quality evidence is still limited.

To address this knowledge gap, we analyzed data from a well-characterized, biopsy-confirmed cohort of IgAN patients enrolled in the Japan IgA Nephropathy Cohort Study (J-IGACS) [11, 12]. We aimed to investigate the association between the magnitude of proteinuria reduction at 12 months post-diagnosis and both a composite kidney outcome and the longitudinal trajectory of eGFR. We further explored whether this association persists across clinically relevant subgroups, including those with characteristics commonly considered in the context of interventional clinical trial eligibility.

## Methods

### Study population

This study represents a *post-hoc* analysis of the Japan IgA Nephropathy Cohort Study (J-IGACS), a prospective, multicenter registry enrolling patients with primary IgAN across Japan between April 1, 2005, and August 31, 2015 [11, 12]. Eligibility required a biopsy-confirmed diagnosis of IgAN and a kidney biopsy specimen containing at least 10 glomeruli for primary study. The study population consisted of individuals from the primary cohort after excluding those with missing values in any covariates included in the multivariable adjustment. Ethical approval was granted by The Jikei University School of Medicine (approval no. 37-069 [12706]). This study was conducted in accordance with the Declaration of Helsinki and reported in compliance with STROBE guidelines.

### Data collection and definitions

Clinical data were collected at the time of biopsy and subsequently at 6-month intervals, including age, sex, blood pressure, serum creatinine, estimated glomerular filtration rate (eGFR), serum uric acid, 24-hour urinary protein excretion (UPE) (the urinary protein-to-creatinine ratio [UPCR] was permitted in place of 24-hour urinary protein excretion at all time points except baseline), and urinary red blood cell count. eGFR was calculated using Uemura’s equation [13] for individuals under 20 years of age and Matsuo’s formula [14] thereafter. Treatments initiated within 1 year after biopsy were considered initial interventions and included corticosteroid therapy, tonsillectomy, and renin-angiotensin-aldosterone system (RAAS) inhibitor use, including any prescriptions ongoing at the time of biopsy. Kidney biopsy histopathological findings were scored using the Oxford MEST-C classification, based on standardized definitions for mesangial (M) and endocapillary (E) hypercellularity, segmental sclerosis (S), tubular atrophy/interstitial fibrosis (T), and crescents (C), in accordance with prior literature [5, 6].

### Exposure and outcome measures

Proteinuria reduction was assessed using the proteinuria ratio, defined as the ratio of proteinuria at 12 months to baseline proteinuria. It was categorized into tertiles or log-transformed for analyses as needed. The primary endpoint was defined as either a ≥40% decrease in eGFR or the initiation of kidney replacement therapy.

### Statistical analysis

Baseline patient characteristics were presented as mean (standard deviation) or median [interquartile range] for continuous variables, and as frequencies and proportions for categorical variables. The Jonckheere–Terpstra test and Cochran–Armitage test were used to evaluate trends in baseline characteristics across tertiles of the proteinuria ratio.

For kidney outcomes, the cumulative incidence of the composite endpoint, defined as a 40% decline in eGFR or initiation of kidney replacement therapy, was estimated using Kaplan–Meier survival curves with the log-rank test for trend across tertiles of proteinuria ratio. Cox proportional hazards models were used to estimate hazard ratios (HRs) and 95% confidence intervals (CIs) for the composite endpoint. These models were adjusted for established risk factors, namely, age, sex, systolic blood pressure, baseline eGFR, baseline log-transformed proteinuria, Oxford MEST-C scores, tonsillectomy, and initiation of RAAS inhibitors and corticosteroids within one year after diagnosis. In addition, to explore the association of continuous values of proteinuria ratio to HR of the composite endpoint, spline regression models were also performed with the median value of the lowest tertile as the reference point and knots at the 5th, 35th, 65th, and 95th percentiles of the proteinuria ratio distribution. Additional analyses were conducted by restricting the study population to a subgroup with baseline eGFR ≥30 and <120 mL/min/1.73 m^2^ and baseline proteinuria ≥0.5 and <3.5 g/day, a range often considered clinically relevant in therapeutic research, to evaluate the robustness of the main findings.

To assess longitudinal changes in kidney function, a linear mixed-effects model for repeated measures was employed to estimate the annual eGFR slope based on the relationship between follow-up time and eGFR levels, where a variance components matrix for the random intercepts to account for individual variability and a first-order autoregressive covariance structure for modeling within-subject correlation across repeated measures were employed. The linearity between follow-up time and eGFR was visually validated using a generalized additive model with smoothing functions.

A sensitivity analysis was performed using the proteinuria ratio at 6 months, a shorter time period compared to the main analysis timeframe, as the explanatory variable.

A two-sided P-value <0.05 was considered statistically significant. All statistical analyses, except for generalized additive model fitting, were performed using SAS version 9.4 (SAS Institute Inc., Cary, NC, USA). Generalized additive model analyses were conducted using R version 4.5.0 (R Foundation for Statistical Computing, Vienna, Austria).

## Results

### Baseline characteristics and follow-up

After excluding individuals with missing values in any of the covariates from the original cohort of 991 patients, a total of 793 patients with complete baseline variables were included in the analysis. The proteinuria ratio, defined as the ratio of proteinuria at 12 months to baseline proteinuria, was categorized into tertiles (Tertile 1, <0.194; Tertile 2, 0.194–0.625; and Tertile 3, ≥0.625). Baseline characteristics showed that the proteinuria ratio was significantly associated with several clinical parameters, including urinary protein excretion, microscopic hematuria, the presence of E, S, and C lesions according to the Oxford classification, initiation of steroid therapy within 1 year, and tonsillectomy within 1 year.

### Association between initial proteinuria reduction and primary outcome

Kaplan–Meier analysis was used to analyze the associations between the kidney composite endpoint, which was defined as a 40% decline in eGFR or initiation of kidney replacement therapy, and the proteinuria ratio. The analysis demonstrated that higher proteinuria ratios were significantly associated with an increased cumulative incidence of the composite endpoint, as indicated by a significant trend in the log-rank test (P for trend <0.001; **Figure 1**). Moreover, in multivariable Cox proportional hazards models adjusted for age, sex, systolic blood pressure, Oxford MEST-C scores, baseline eGFR, baseline log-transformed proteinuria, tonsillectomy, initiation of RAAS inhibitor therapy, and initiation of steroid therapy, higher proteinuria ratios remained independently associated with an elevated risk of the incident composite endpoint (**Table 2**). Additional analyses using spline models, with the median value of Tertile 1 used as the reference point, revealed that the risk of the incident composite endpoint increased continuously as the proteinuria ratio rose above the median value (**Figure 2**).

**Figure 1.**
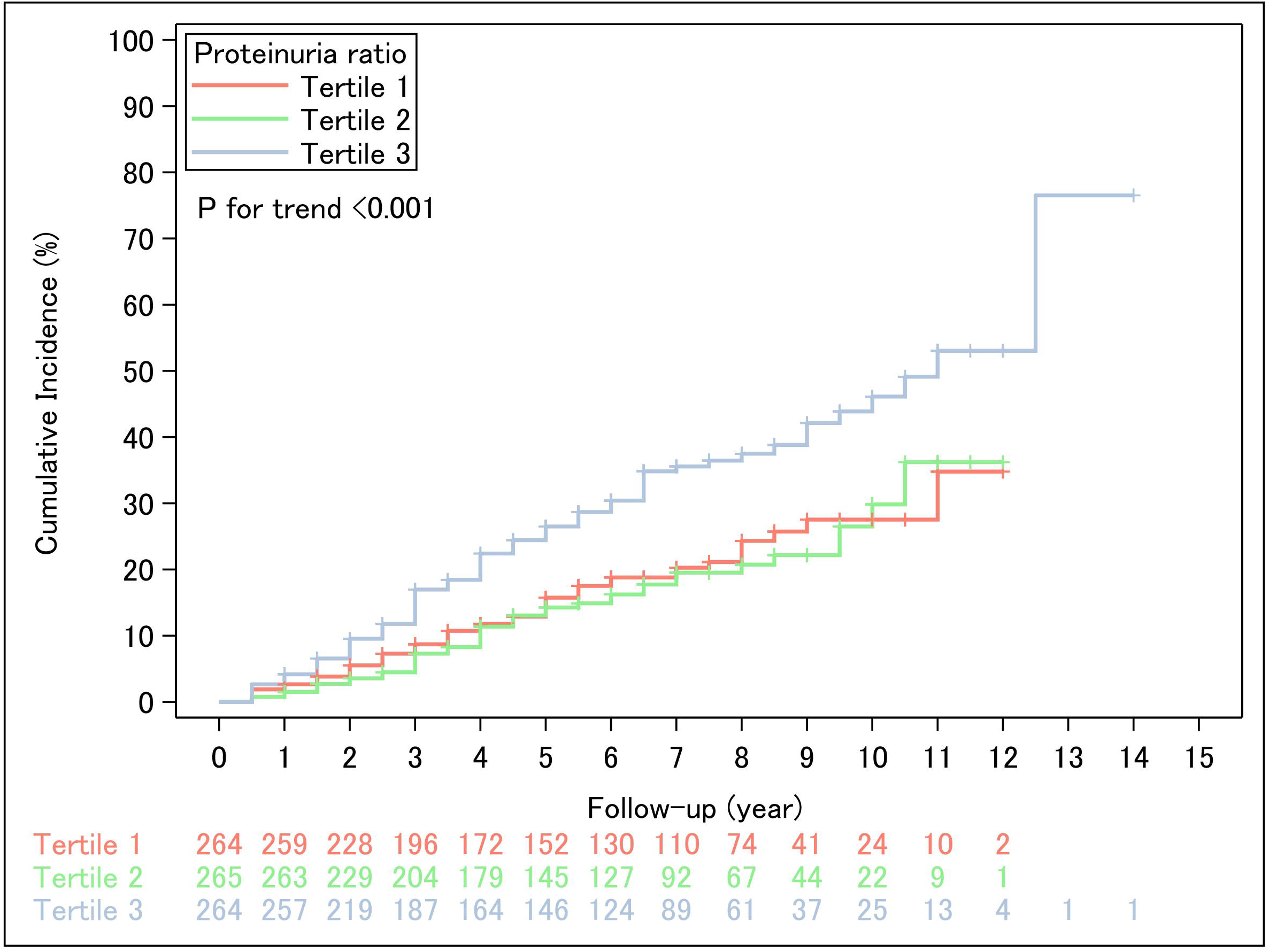
Kaplan–Meier curves for the kidney composite endpoint according to proteinuria ratio tertiles. Kaplan–Meier survival curves show the cumulative incidence of the kidney composite endpoint, defined as a 40% decline in estimated glomerular filtration rate or initiation of kidney replacement therapy, stratified by tertiles of the proteinuria ratio at 12 months (Tertile 1,<0.194; Tertile 2, 0.194–0.625; Tertile 3, ≥0.625).

**Figure 2.**
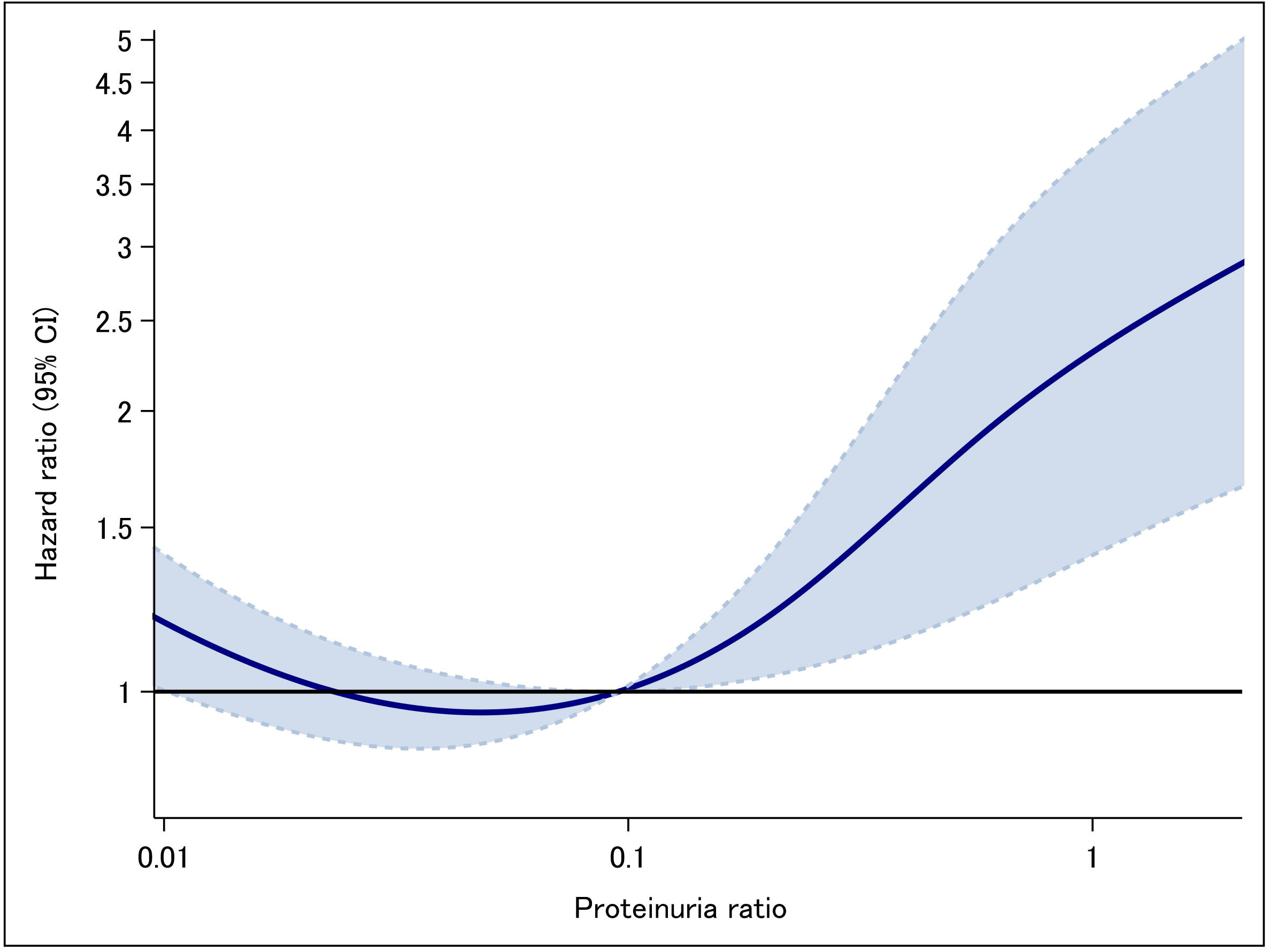
Adjusted hazard ratios for the kidney composite endpoint by proteinuria ratio tertiles. Forest plot displaying adjusted hazard ratios for the kidney composite endpoint according to proteinuria ratio tertiles, using a multivariable Cox proportional hazards model. Adjustment variables included age, sex, systolic blood pressure, initiation of renin-angiotensin-aldosterone inhibitor therapy, initiation of steroid therapy, tonsillectomy, Oxford MEST-C* scores, baseline estimated glomerular filtration rate, and log-transformed proteinuria. The risk of adverse outcomes increased with higher proteinuria ratios. *Oxford MEST-C indicates mesangial hypercellularity, endocapillary hypercellularity, segmental sclerosis, tubular atrophy/interstitial fibrosis, and cellular/fibrocellular crescents.

**Table 1.** Baseline characteristics and initial treatment according to proteinuria ratio tertiles.

|  | Proteinuria ratio |  |  | P for trend |
| --- | --- | --- | --- | --- |
|  | Tertile 1 | Tertile 2 | Tertile 3 |  |
|  | (<0.194)<br>n = 264 | (0.194–0.625)<br>n = 265 | (≥0.625)<br>n = 264 |  |
| **Clinical characteristics** |  |  |  |  |
| Age, year | 37.9 (16.6) | 38.9 (16.0) | 40.2 (17.0) | 0.13 |
| Male sex, n (%) | 140 (53.0) | 138 (52.1) | 129 (48.5) | 0.30 |
| Height, cm | 161.2 (12.5) | 161.9 (11.0) | 162.4 (10.1) | 0.54 |
| Weight, kg | 58.5 (14.5) | 59.1 (13.9) | 59.1 (12.6) | 0.70 |
| Systolic blood pressure, mmHg | 122.0 (18.0) | 121.4 (17.4) | 123.2 (17.7) | 0.37 |
| eGFR, mL/min/1.73 m<sup>2</sup> | 75.5 (29.2) | 75.4 (26.9) | 76.3 (29.4) | 0.46 |
| UPE, g/day | 1.1 [0.5–2.1] | 0.6 [0.4–1.1] | 0.4 [0.2–0.8] | <0.001 |
| Microhematuria*, n (%) | 239 (90.5) | 226 (85.3) | 208 (78.8) | <0.001 |
| **Pathological characteristics†** |  |  |  |  |
| M1, n (%) | 93 (35.2) | 70 (26.4) | 68 (25.8) | 0.017 |
| E1, n (%) | 125 (47.4) | 94 (35.5) | 69 (26.1) | <0.001 |
| S1, n (%) | 216 (81.8) | 203 (76.6) | 177 (67.1) | <0.001 |
| T1 or T2, n (%) | 62 (23.5) | 50 (19.3) | 65 (24.2) | 0.83 |
| C1 or C2, n (%) | 150 (56.8) | 94 (35.5) | 67 (25.4) | <0.001 |
| **Initial treatment** |  |  |  |  |
| RAAS inhibitor, n (%) | 155 (58.7) | 154 (58.1) | 165 (62.5) | 0.37 |
| Corticosteroid, n (%) | 222 (84.1) | 190 (71.7) | 114 (42.8) | <0.001 |
| Tonsillectomy, n (%) | 138 (39.4) | 121 (45.7) | 91 (34.5) | <0.001 |

Abbreviations: eGFR, estimated glomerular filtration rate; RAAS, renin-angiotensin-aldosterone system; UPE, urinary protein excretion.
\*Defined as 5 or more red blood cells / high-power field
†Indicates Oxford MEST-C scores: M, mesangial hypercellularity; E, endocapillary hypercellularity; S, segmental sclerosis; T, tubular atrophy/interstitial fibrosis; and C, cellular/fibrocellular crescents.

**Table 2.** Hazard ratios (HRs) for kidney composite endpoint according to proteinuria ratio tertiles.

| Tertile | No. of<br>outcome<br>s | No. of<br>participant<br>s | Age- and<br>sex-adjusted |  | Multivariable<br>adjusted* |  |
| --- | --- | --- | --- | --- | --- | --- |
|  |  |  | HR (95%<br>CI) | P for<br>trend | HR (95%<br>CI) | P for<br>trend |
| Tertile 1<br>($<0.194$) | 48 | 264 | 1 (ref) | | 1 (ref) | |
| Tertile 2<br>($0.194\text{--}0.625$) | 43 | 265 | 0.93<br>(0.61–1.40) | $<0.001$ | 1.00<br>(0.65–1.54) | 0.021 |
| Tertile 3<br>($\geq 0.625$) | 85 | 264 | 1.65<br>(1.30–2.63) | | 1.65<br>(1.09–2.52) | |

Abbreviation: CI, confidence interval.
\*Adjusted for age, sex, systolic blood pressure, baseline estimated glomerular filtration rate, baseline log-transformed proteinuria, and Oxford MEST-C scores<sup>†</sup>, initiation of renin-angiotensin-aldosterone system inhibitors and corticosteroids within one year after diagnosis, tonsillectomy.
<sup>†</sup>Oxford MEST-C indicates mesangial hypercellularity, endocapillary hypercellularity, segmental sclerosis, tubular atrophy/interstitial fibrosis, cellular/fibrocellular crescents.

### Restricted cohort analysis

To further assess the robustness of the findings, we performed an analysis limited to patients with baseline eGFR ≥30 and <120 mL/min/1.73 m^2^ and baseline proteinuria ≥0.5 and <3.5 g/day, a range often considered relevant in clinical and research contexts. These analyses demonstrated results consistent with the main findings, and the association between a higher proteinuria ratio and adverse kidney outcomes appeared even stronger within this restricted cohort (**Figure 3**).

**Figure 3.**
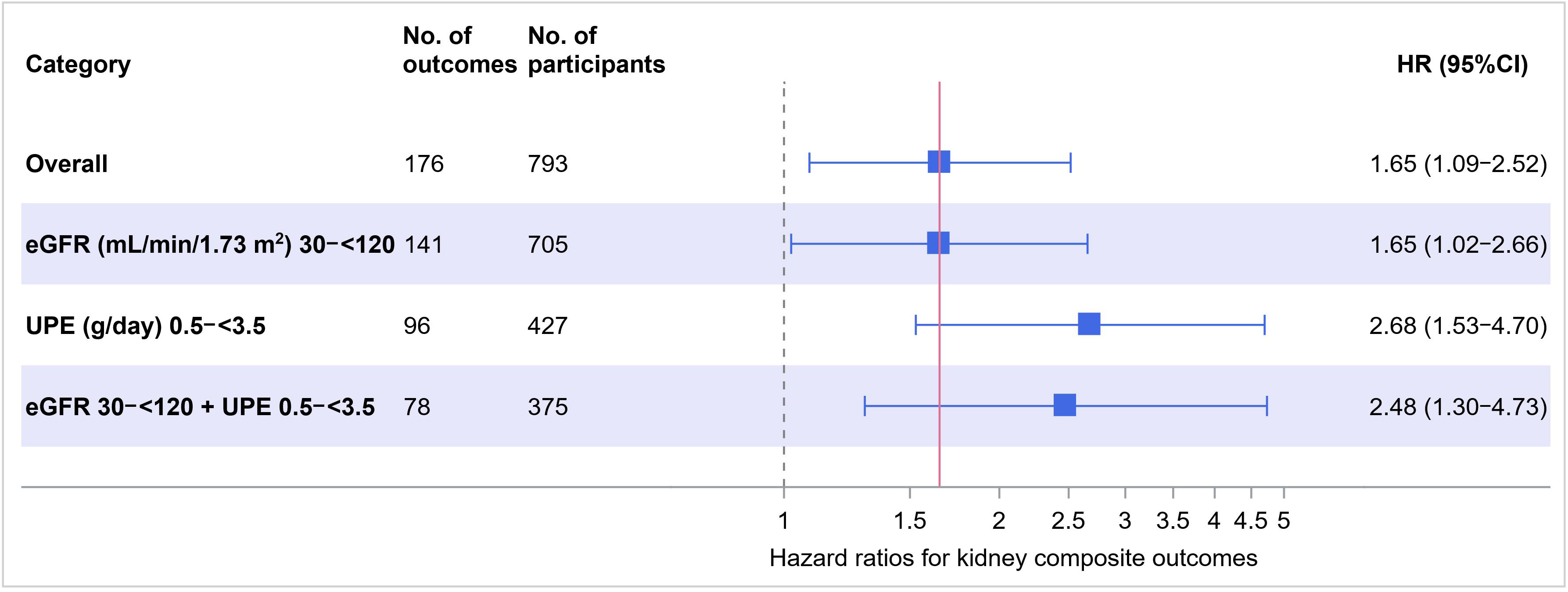
Association between continuous proteinuria ratio and hazard ratio (HR) of the kidney composite endpoint using spline models. Spline curves depict the continuous relationship between the proteinuria ratio at 12 months and the multivariable-adjusted HR for the kidney composite endpoint, where the median proteinuria ratio of the 1^st^ tertile was treated as the reference point and the 5th, 35th, 65th, and 95^th^ points were used as knots. The model was adjusted for age, sex, systolic blood pressure, and initiation of renin-angiotensin-aldosterone system inhibitors or corticosteroids within one year after diagnosis, tonsillectomy, baseline estimated glomerular filtration rate, baseline log-transformed proteinuria, and Oxford MEST-C scores*. *Oxford MEST-C indicates mesangial hypercellularity, endocapillary hypercellularity, segmental sclerosis, tubular atrophy/interstitial fibrosis, and cellular/fibrocellular crescents.

### Association of initial proteinuria reduction and eGFR slope

A multivariable-adjusted mixed-effects model for repeated measures demonstrated that higher proteinuria ratios were significantly associated with a steeper decline in eGFR slope (P for trend <0.001; **Figure 4**).

**Figure 4.**
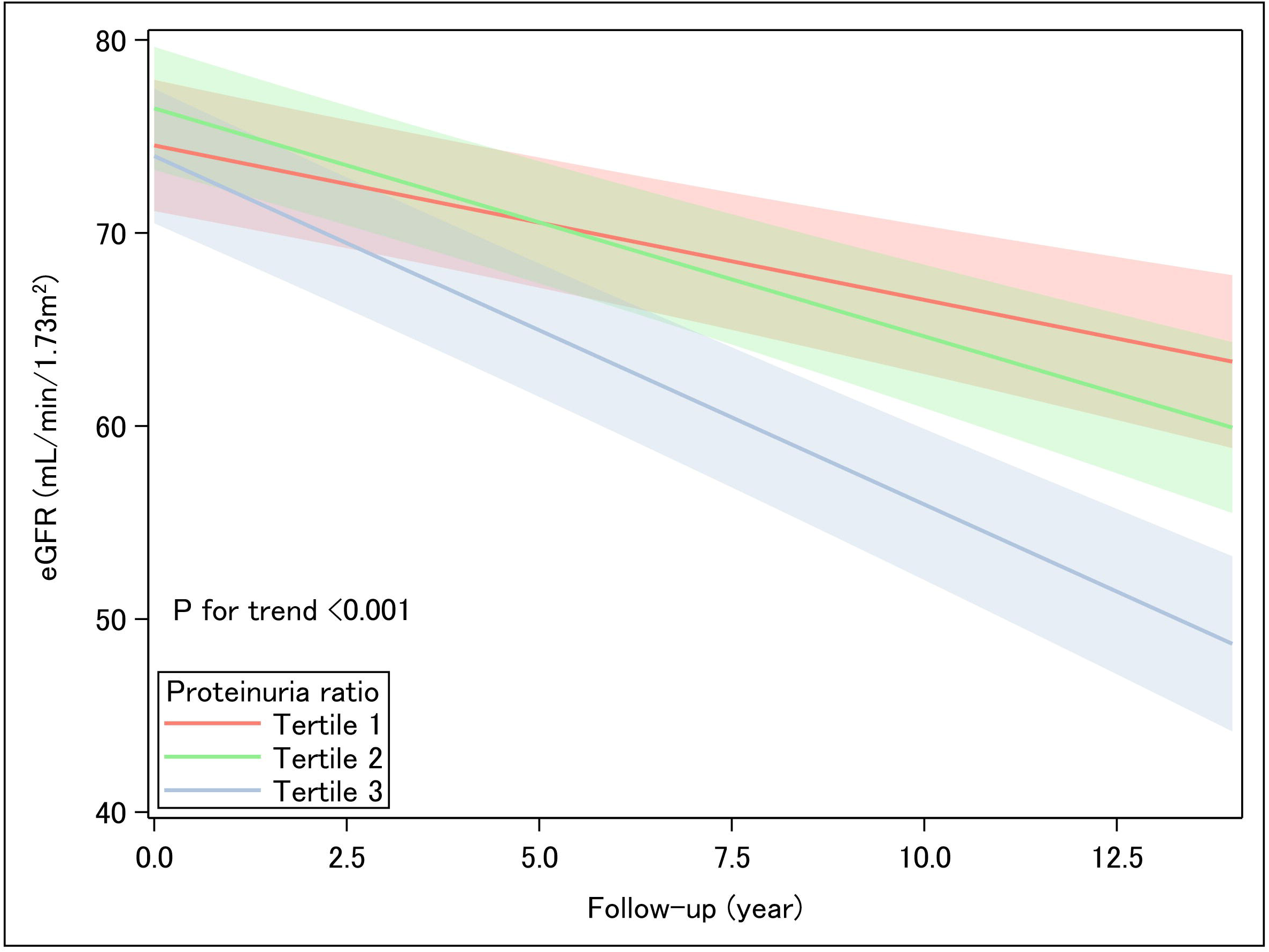
Annual slope of the estimated glomerular filtration rate (eGFR) according to proteinuria ratio tertiles. The association between eGFR levels and follow-up time was analyzed using a mixed-effects model for repeated measures across proteinuria ratio tertiles (Tertile 1, <0.194; Tertile 2, 0.194–0.625; Tertile 3, ≥0.625). The model was adjusted for age, sex, systolic blood pressure, and initiation of renin-angiotensin-aldosterone system inhibitors or corticosteroids within one year after diagnosis, tonsillectomy, baseline estimated glomerular filtration rate, baseline log-transformed proteinuria, and Oxford MEST-C scores*. * Oxford MEST-C indicates mesangial hypercellularity, endocapillary hypercellularity, segmental sclerosis, tubular atrophy/interstitial fibrosis, and cellular/fibrocellular crescents.

### Sensitivity analysis

As a sensitivity analysis, we conducted an additional analysis using the proteinuria ratio at 6 months, a shorter time period compared to the main analysis timeframe, as an explanatory variable. Both multivariable Cox proportional hazards models and mixed-effects models for repeated measures yielded significant associations consistent with the primary analyses, although the associations were attenuated (**Supplementary Table S1, Supplementary Figure S1**).

## Discussion

In this post hoc analysis of a large-scale, nationwide, prospective cohort of biopsy-confirmed IgAN patients, we found that early reduction in proteinuria, quantified at 12 months after histopathological diagnosis using the proteinuria ratio, was independently and consistently associated with the long-term clinical endpoints, defined as a 40% or greater decline in eGFR or initiation of kidney replacement therapy. Moreover, early reduction in proteinuria was found to be consistently associated with the intermediate-term trajectories of kidney function decline assessed by the eGFR slope. The latter association remained significant even after multivariable adjustment for demographic, clinical, and histopathological covariates, and across sensitivity analyses. These results reinforce the association between early proteinuria response and long-term kidney outcomes, highlighting the potential of early proteinuria response as a clinically actionable, pathophysiologically relevant, and dynamically measurable surrogate endpoint. Incorporating proteinuria reduction into risk stratification algorithms and treatment response evaluation may enhance individualized clinical decision-making in the management of IgAN.

From a mechanistic perspective, proteinuria serves as a surrogate indicator of structural injury to the glomerular filtration barrier and reflects ongoing immunologically mediated inflammation [15, 16]. A reduction in proteinuria, therefore, is likely indicative of favorable modulation of underlying pathogenic pathways. Importantly, proteinuria possesses several practical advantages over other biomarkers: it is routinely quantified in clinical practice, exhibits prompt responsiveness to therapeutic intervention, and is readily interpretable at the point of care. These attributes collectively render it highly amenable to incorporation into dynamic, real-time clinical decision-making frameworks. Our data provide compelling evidence that, even within the constraints of a real-world, non-randomized setting, early amelioration of proteinuria is independently associated with long-term kidney function preservation [7]. In addition to its established association with definitive clinical endpoints, our analysis revealed that a lower proteinuria ratio was significantly correlated with a more gradual subsequent decline in eGFR, consistent with previous meta-regression analyses that evaluated the relationship between treatment-induced reductions in proteinuria and improvements in eGFR decline [17, 18]. This observation supports the hypothesis that proteinuria may function as an upstream surrogate marker, reflecting both functional impairment and structural progression of kidney disease [19, 20]. These findings lend further biological and clinical plausibility to the use of proteinuria as a comprehensive indicator of disease activity and therapeutic response in IgAN.

From a clinical perspective, these findings offer practical and actionable insights for the early stratification of patient risk in IgAN [7, 15, 21, 22]. Individuals who achieve a marked reduction in proteinuria within the first year following diagnosis may be identified as having a more favorable prognosis. This subgroup may be appropriate for de-escalation of immunosuppressive regimens, reduction in monitoring frequency, and provision of reassurance based on observed associations with favorable outcomes. In contrast, patients demonstrating insufficient proteinuria reduction may warrant intensified therapeutic strategies, closer clinical surveillance, or consideration for inclusion in interventional clinical trials. Owing to its therapeutic modifiability, ease of measurement, and interpretability in routine practice, proteinuria represents a particularly valuable parameter for dynamic risk assessment and response-adaptive treatment strategies, principles that lie at the core of precision nephrology.

To further validate the clinical applicability of our findings, we conducted an additional analysis restricted to patients with baseline proteinuria between 0.5 and 3.5 g/day, a range often considered clinically relevant in therapeutic contexts [23]. This subgroup reflects a population at moderate to high risk, in whom treatment decisions are frequently guided by proteinuria levels [24]. Additionally, we performed a subgroup analysis limited to patients with baseline eGFR between 30 and 120 mL/min/1.73 m^2^, representing a population for whom kidney function is preserved enough to permit intervention. The consistent and even strengthened associations observed in both subgroups support the robustness of proteinuria reduction as a meaningful marker and highlight its significance for clinical trial design and real-world decision-making in IgAN.

Several limitations of this study should be acknowledged. First, although the cohort was prospectively assembled, this was a *post hoc* analysis and is therefore subject to inherent limitations of observational research, including potential residual confounding. Second, proteinuria measurements were obtained according to institutional protocols rather than centralized standardization, potentially introducing inter-site variability. Third, the 12-month window used for assessing proteinuria reduction, while clinically pragmatic, may not fully capture heterogeneity in early treatment responses [25]. However, our sensitivity analysis using the 6-month proteinuria ratio yielded consistent results, suggesting that evaluation at 6 to 12 months may be acceptable in clinical and research settings. Lastly, our cohort consisted exclusively of Japanese patients. While this homogeneity offers unique insights into a high-prevalence population [26], it may limit the generalizability of our findings to other ethnic and geographic groups with differing genetic predispositions, environmental exposures, and healthcare systems.

In conclusion, early reduction in proteinuria serves as a reliable, pathophysiologically meaningful, and clinically applicable marker associated with both long-term and intermediate kidney outcomes in patients with IgAN. Its availability, responsiveness to therapy, and consistent association with clinical outcomes highlight its value as a surrogate endpoint in both clinical practice and clinical research. Future studies should aim to refine optimal thresholds, examine disease- and treatment-specific responsiveness, and validate these findings across more diverse populations and settings.

## Authors’ Contributions

Conceptualization: Takaya Sasaki, Nobuo Tsuboi, Takashi Yokoo, Yusuke Suzuki.

Data curation: Takaya Sasaki.

Formal analysis: Takaya Sasaki.

Funding acquisition: Yusuke Suzuki.

Investigation: Takaya Sasaki, Nobuo Tsuboi.

Methodology: Takaya Sasaki.

Project administration: Nobuo Tsuboi, Takashi Yokoo, Yusuke Suzuki.

Supervision: Takashi Yokoo, Yusuke Suzuki.

Visualization: Takaya Sasaki.

Writing – original draft: Takaya Sasaki, Nobuo Tsuboi.

Writing – review & editing: Kentaro Koike, Hiroyuki Ueda, Masahiro Okabe, Shinya Yokote, Akihiro Shimizu, Keita Hirano, Tetsuya Kawamura, Takashi Yokoo, Yusuke Suzuki.

## Funding

This study was partly supported by a Grant-in-Aid for Progressive Renal Diseases Research, Research on Rare and Intractable Disease, from the Ministry of Health, Labour and Welfare of Japan. This research was supported by the Japan Agency for Medical Research and Development under grant JP19ek0109261.

## Financial Disclosure

YS has received consulting fees from Otsuka Pharmaceutical (Visterra), Novartis, Chinook Therapeutics, ARGENX, BioCryst, Alexion Pharmaceuticals, Renalys, Alpine, and George Clinical.

YS has also received honoraria for lectures, presentations, manuscript writing, or educational events from Kyowa Kirin, Novartis, Mitsubishi Tanabe, Otsuka Pharmaceutical, Daiichi Sankyo, AstraZeneca, Boehringer Ingelheim, and Chinook Therapeutics.

### Acknowledgments

This study was approved by the ethics review board of the Jikei University School of Medicine (37-069[12706]).

## Data availability statement

All data produced in the present study are available upon reasonable request to the authors.

## Notes

### Summary of Updates

Minor title change. Updated COI disclosure.

