## Supplemental materials for "Initial proteinuria reduction and adverse kidney outcomes in IgA nephropathy: An analysis from the J-IGACS"

**Table of Contents**

**Investigators list of the J-IGACS working group.**
**Supplementary Table S1. Hazard ratios (HRs) for the kidney composite endpoint by tertiles of the proteinuria ratio at 6 months.**
**Supplementary Figure S1. Annual slope of the estimated glomerular filtration rate (eGFR) by tertiles of the proteinuria ratio at 6 months.**

**Investigators list of the J-IGACS working group.**
**Chair:** Yusuke Suzuki (Department of Nephrology, Juntendo University Faculty of Medicine, Tokyo, Japan.)
**Co-chair:** Takashi Yokoo (Division of Nephrology and Hypertension, Department of Internal Medicine, The Jikei University School of Medicine, Tokyo, Japan.)

**Investigators:**
Ryosuke Aoki (Department of Nephrology, Juntendo University Faculty of Medicine, Tokyo, Japan.)
Shouichi Fujimoto (Department of Medical Environment Innovation, Faculty of Medicine, University of Miyazaki, Miyazaki, Japan.)
Yusuke Fukao (Department of Nephrology, Juntendo University Faculty of Medicine, Tokyo, Japan.)
Akihiro Fukuda (Department of Endocrinology, Metabolism, Rheumatology and Nephrology, Faculty of Medicine, Oita University, Oita, Japan.)
Akinori Hashiguchi (Department of Pathology, Keio University School of Medicine, Tokyo, Japan.)
Hiroshi Hataya (Department of Nephrology and Rheumatology, Tokyo Metropolitan Children's Medical Center, Fuchu, Tokyo, Japan.)
Keita Hirano (Division of Nephrology and Hypertension, Department of Internal Medicine, The Jikei University School of Medicine, Tokyo, Japan.)
Shiko Honma (Department of Pathology, The Jikei University School of Medicine, Tokyo, Japan.)
Daisuke Ichikawa (Division of Nephrology and Hypertension, Department of Internal Medicine, St. Marianna University School of Medicine, Kanagawa, Japan.)
Takafumi Ito (Department of Internal Medicine, Nephrology, Teikyo University School of Medicine, Teikyo University Chiba Medical Center, Chiba, Japan.)
Kensuke Joh (Department of Pathology, The Jikei University School of Medicine, Tokyo, Japan.)
Ritsuko Katafuchi (Kidney Unit, National Hospital Organization, Fukuoka-Higashi Medical Center, Fukuoka, Japan. Division of Nephrology, Medical Corporation Houshikai, Kano Hospital.)
Tetsuya Kawamura (Division of Nephrology and Hypertension, Department of Internal Medicine, The Jikei University School of Medicine, Tokyo, Japan.)
Masao Kihara (Department of Nephrology, Juntendo University Faculty of Medicine, Tokyo, Japan.)
Masao Kikuchi (Division of Cardiovascular Medicine and Nephrology, Department of Internal Medicine, Faculty of Medicine, University of Miyazaki, Miyazaki, Japan.)
Kentaro Koike (Division of Nephrology and Hypertension, Department of Internal Medicine, The Jikei University School of Medicine, Tokyo, Japan.)
Keiichi Matsuzaki (Department of Public Health, Kitasato University School of Medicine, Kanagawa, Japan.)
Kenichiro Miura (Department of Pediatric Nephrology, Tokyo Women's Medical University, Tokyo, Japan.)
Yoichi Miyazaki (Division of Nephrology and Hypertension, Department of Internal Medicine, The Jikei University School of Medicine, Tokyo, Japan.)
Takahito Moriyama (Department of Nephrology, Tokyo Medical University, Tokyo, Japan.)
Kumiko Muta (Advanced Medical Education Center, Nagasaki University School of Medicine, Nagasaki, Japan.)

Koichi Nakanishi (Department of Child Health and Welfare (Pediatrics), Graduate School of Medicine, University of the Ryukyus, Ginowan, Okinawa, Japan.)
Shinya Nakatani (Department of Metabolism, Endocrinology and Molecular Medicine, Osaka Metropolitan University Graduate School of Medicine, Osaka, Japan.)
Yoshihito Nihei (Department of Nephrology, Juntendo University Faculty of Medicine, Tokyo, Japan.)
Masako Nishikawa (Center for Reseach Promotion, The Jikei University School of Medicine, Tokyo, Japan.)
Tomoya Nishino (Department of Nephrology, Graduate School of Biomedical Sciences, Nagasaki University, Nagasaki, Japan.)
Ryoko Sakaguchi (Department of Pathology, The Jikei University School of Medicine, Tokyo, Japan.)
Takaya Sasaki (Division of Nephrology and Hypertension, Department of Internal Medicine, The Jikei University School of Medicine, Tokyo, Japan.)
Satoru Sanada (Department of Nephrology, Japan Community Healthcare Organization Sendai Hospital, Sendai, Japan.)
Sayuri Shirai (Division of Nephrology and Hypertension, Department of Internal Medicine, St. Marianna University School of Medicine, Kanagawa, Japan.)
Akihiro Shimizu (Division of Nephrology and Hypertension, Department of Internal Medicine, The Jikei University School of Medicine, Tokyo, Japan.)
Akira Shimizu (Department of Analytic Human Pathology, Nippon Medical School, Tokyo, Japan.)
Takanori Shibata (Division of Nephrology, Department of Medicine, Showa Medical University School of Medicine, Tokyo, Japan.)
Yuko Shima (Department of Pediatrics, Wakayama Medical University, Wakayama City, Wakayama, Japan.)
Hitoshi Suzuki (Department of Nephrology, Juntendo University Faculty of Medicine, Tokyo, Japan.)
Kazuo Takahashi (Department of Biomedical Molecular Sciences, School of Medicine, Fujita Health University, Nagoya, Aichi, Japan.)
Nobuo Tsuboi (Division of Nephrology and Hypertension, Department of Internal Medicine, The Jikei University School of Medicine, Tokyo, Japan.)
Yasuhiko Tomino (Asian Pacific Renal Research Promotion Office, Medical Corporation SHOWAKAI, Shinjuku-ku, Tokyo, Japan.)
Hiroyuki Ueda (Division of Nephrology and Hypertension, Department of Internal Medicine, The Jikei University School of Medicine, Tokyo, Japan.)
Maki Urushihara (Department of Pediatrics, Institute of Biomedical Sciences, Tokushima University Graduate School, Tokushima, Tokushima, Japan.)
Takashi Yasuda (Naruse Kidney Clinic, Tokyo, Japan.)
Yoshinari Yasuda (Department of Advanced Science in Renal-Cardio Medicine/Nephrology, Gifu University Graduate School of Medicine, Gifu, Japan.)
Shinya Yokote (Department of Nephrology, Kawaguchi Municipal Medical Center, Saitama, Japan.)

**Supplementary Table S1. Hazard ratios (HRs) for the kidney composite endpoint by tertiles of the proteinuria ratio at 6 months.**

| Tertile | No. of outcomes | No. of participants | Age- and sex-adjusted | | Multivariable adjusted* | |
|---|---|---|---|---|---|---|
| | | | HR (95% CI) | P for trend | HR (95% CI) | P for trend |
| Tertile 1 (<0.326) | 55 | 276 | 1 (ref) | | 1 (ref) | |
| Tertile 2 (0.326–4.78) | 58 | 276 | 1.04 (0.72–1.99) | 0.034 | 1.06 (0.72–2.02) | 0.036 |
| Tertile 3 (≥4.78) | 78 | 276 | 1.44 (1.02–1.39) | | 1.49 (1.01–1.45) | |

Abbreviation: CI, confidence interval.
*Adjusted for age, sex, systolic blood pressure, baseline estimated glomerular filtration rate, baseline log-transformed proteinuria, Oxford MEST-C scores†, initiation of renin-angiotensin-aldosterone system inhibitors and corticosteroids within one year after diagnosis, and tonsillectomy.
†Oxford MEST-C indicates mesangial hypercellularity, endocapillary hypercellularity, segmental sclerosis, tubular atrophy/interstitial fibrosis, and cellular/fibrocellular crescents.

**Supplementary Figure S1. Annual slope of the estimated glomerular filtration rate (eGFR) by tertiles of the proteinuria ratio at 6 months.**

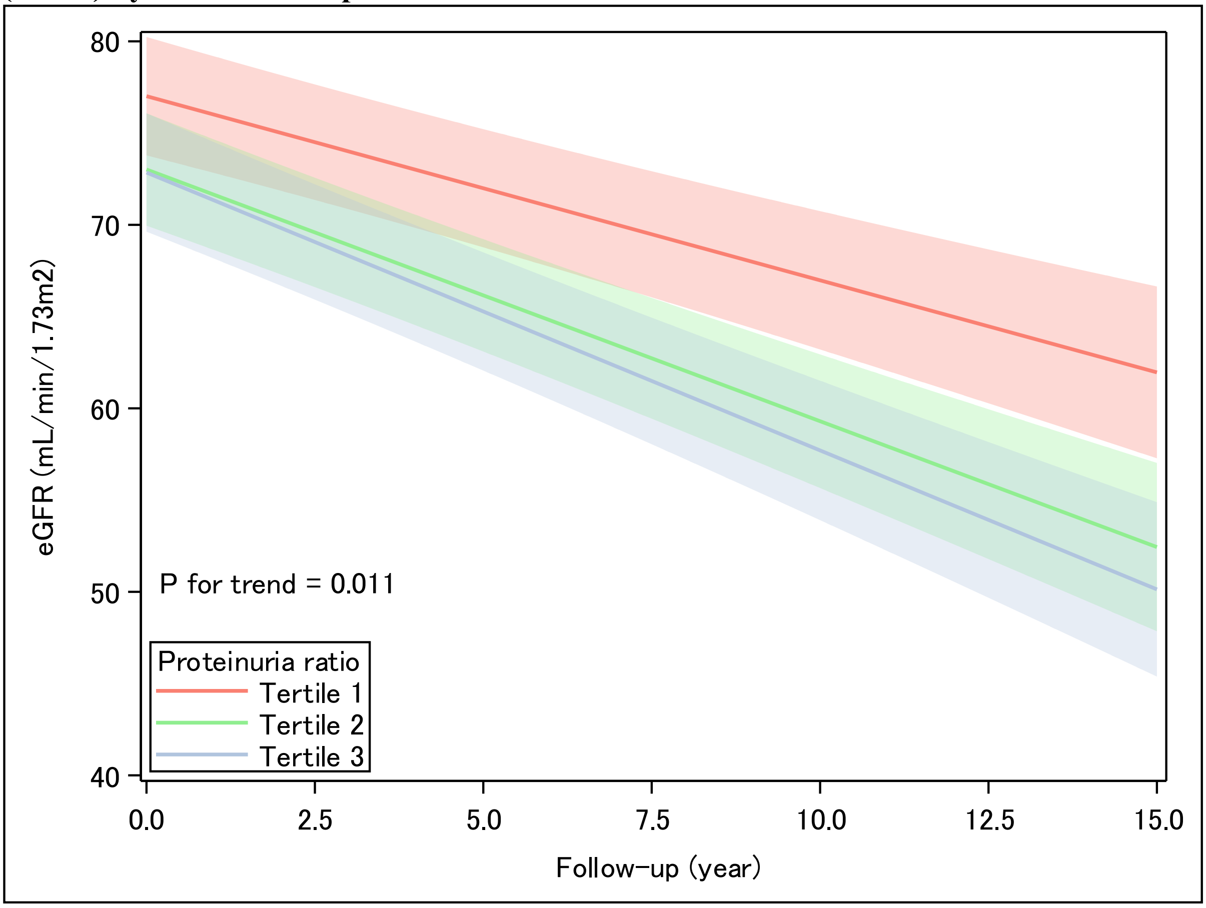


The association between eGFR levels and follow-up time was analyzed using a mixed-effects model for repeated measures across tertiles of the proteinuria ratio at 6 months (Tertile 1, <0.326; Tertile 2, 0.326–4.78; Tertile 3, ≥4.78). The model was adjusted for age, sex, systolic blood pressure, initiation of renin-angiotensin-aldosterone system inhibitors and corticosteroids within one year after diagnosis, tonsillectomy, baseline estimated glomerular filtration rate, baseline log-transformed proteinuria, and Oxford MEST-C scores*.
*Oxford MEST-C indicates mesangial hypercellularity, endocapillary hypercellularity, segmental sclerosis, tubular atrophy/interstitial fibrosis, and cellular/fibrocellular crescents.
